# Individualised Functional Brain Mapping Distinguishes Drug-Resistant from Early-Stage Epilepsy

**DOI:** 10.64898/2026.02.12.26346195

**Authors:** Mangor Pedersen, Donna Parker, Graeme D. Jackson, the Australian Epilepsy Project Investigators

**Affiliations:** Department of Psychology and Neuroscience, Auckland University of Technology, Auckland, New Zealand; The Florey Institute of Neuroscience and Mental Health, Melbourne, Victoria, Australia; Department of Medicine, Austin Health, The University of Melbourne, Australia; Department of Neurology, Austin Health, Melbourne, Australia

## Abstract

Epilepsy is among the most prevalent neurological disorders, affecting millions of individuals worldwide at every stage of life. Characterised by recurrent seizures, epilepsy can significantly disrupt daily functioning, education, employment, and overall quality of life. Despite advances in neuroimaging, current approaches often overlook the individualised nature of brain disruptions in epilepsy. Here, we introduce an individualised functional Magnetic Resonance Imaging (fMRI) framework, Adjusted Local Estimates of Connectivity (ALEC), to detect patient-specific brain local connectivity abnormalities across distinct clinical stages of epilepsy. To do so, we analysed movie-watching multi-echo fMRI in 102 heterogeneous epilepsy patients (34 with drug-resistant epilepsy; 34 with a new diagnosis of epilepsy; 34 after a first seizure) and 68 socioeconomically matched healthy controls. ALEC is a voxel-wise modified z-score and estimates deviations in Regional Homogeneity from healthy norms at the individual level. Our results show that whole-brain averaged ALEC scores were significantly higher in drug-resistant epilepsy compared to early-stage cohorts. Several drug-resistant individuals exhibited pronounced ALEC elevations in the hippocampus, thalamus and brainstem alongside widespread cortical decreases, although these patterns did not reach group-level significance. Age and seizure duration correlated positively with ALEC, but only within the drug-resistant group. We also highlight a subset of cases that demonstrated concordance with ALEC and the patient’s clinical history and investigations, including epileptogenic pathology. Combined, our findings highlight the importance of individualised neuroimaging approaches for understanding epilepsy. By revealing biologically concordant local connectivity patterns—marked by local hyperconnectivity in drug-resistant cases worsening with aging—ALEC provides a potential pathway for precision brain mapping of patients at risk for drug resistance.

## Introduction

Epilepsy is a complex neurological disorder characterised by recurrent seizures, with significant variability in clinical presentation, underlying pathology, and treatment response ^1^. This heterogeneity spans seizure types, aetiologies, and comorbidities, making diagnosis and management particularly challenging. While some individuals achieve seizure control with medication, others develop drug-resistant epilepsy, which is associated with increased cognitive, psychological, and social burden ^2^. In routine care, clinicians aim to determine whether a person’s seizures will respond to medication, whether surgery should be considered, and how cognitive and mental health risks should be managed ^3,4^. One of the enduring challenges in epilepsy research and clinical management is early identification of the development of drug resistance, as it can inform treatment strategies and improve prognostic accuracy ^5^.

Recent advances in Functional Magnetic Resonance Imaging (fMRI) and computational modelling offer promising avenues for addressing the diagnostic and prognostic challenges in epilepsy at a group level (e.g., ^6–14^). While group-level designs have been invaluable for establishing disease-specific brain abnormalities, they reveal what is common across patients rather than what is unique to an individual—a limitation that is at odds with the clinical imperatives of precision medicine ^15,16^. A framework for detecting patient-specific functional brain alterations can potentially aid clinical decision-making by capturing subtle, individualised patterns of brain network dysfunction. Such approaches can support early identification of patients at risk for persistent seizures, even in cases where conventional imaging appears normal. Integrating these individualised analyses with clinical and genetic data could further enhance prognostic accuracy and inform the development of tailored treatment strategies.

Recognising the need for tools that capture individual variability in brain network dysfunction, we propose ALEC, a novel fMRI-based method tailored to identify patient-specific connectivity alterations across heterogeneous epilepsy cohorts. ALEC is a voxel-wise modification of fMRI-based Regional Homogeneity ^17,18^, and it aims to quantify individual deviations in functional local connectivity, compared to healthy controls. Recent work has also highlighted the promise of individualised analyses, especially in the normative modelling domain ^19^. For example, Xie et al. ^20^ demonstrated that personalised fMRI biomarkers derived from normative models achieved high accuracy in distinguishing temporal lobe epilepsy from controls, underscoring the clinical potential of individualised neuroimaging markers. Similar non-normative modelling approaches, similar to ALEC, have been used with structural MRI data in various epilepsy subtypes ^21^, as well as Traumatic Brain Injury ^21–23^.

In this study, we analysed data from the Australian Epilepsy Project (https://www.epilepsyproject.org.au/). This large-scale initiative collects multimodal prospective data from individuals with: i) drug-resistant epilepsy (DRE), ii) a recent first seizure (1st Sz), and iii) a recent formal diagnosis of epilepsy (new Dx). In the current study, we leverage these three clinically heterogeneous epilepsy cohorts to seek a mechanistic understanding of epilepsy. The results in this study show that ALEC is elevated in DRE relative to early-stage epilepsy cohorts, most prominently in the thalamus and hippocampus, consistent with their reputed roles in seizure initiation, maintenance, and network synchronisation ^24^. We also show that age correlates with ALEC in drug-resistant epilepsy, potentially reflecting cumulative network reorganisation over disease duration rather than normal brain development ^25^. Lastly, we investigate ALEC on an individual level and show that brain connectivity patterns correspond to clinical investigations, as well as structural MRI and electroencephalogram (EEG) findings.

## Materials and Methods

### Participants and ethics

Participants were drawn from three prospectively defined cohorts from the Australian Epilepsy Project. Cohort assignment followed clinical criteria documented at baseline: DRE was defined by persistent seizures despite adequate trials of anti-seizure medications; 1^st^ Sz participants had unprovoked seizure(s) without prior epilepsy diagnosis; new Dx cases had ≥2 unprovoked seizures within 12 months and/or a diagnosis or epilepsy confirmed by a neurologist. Clinical data were extracted from structured records, including age, gender, MRI findings, EEG findings, and seizure duration (where applicable). The DRE cohort comprised 34 individuals (16/18 M/F; mean age 37.3 ± 13.9 years, range 18–61) with a median seizure duration of 11 years (range 1–59 years). MRI findings in this group included structural abnormalities (24%), prior surgery (18%), incidental findings (12%), and no structural abnormality (38%). EEG was abnormal in most cases, predominantly showing focal epileptiform activity (62%). The 1st Sz cohort included 34 participants (19/15 M/F; mean age 36.2 ± 14.4 years, range 18–66), with MRI largely normal (62%) and EEG predominantly normal (82%). The New Dx cohort comprised 34 participants (20/14 M/F; mean age 31.1 ± 9.5 years, range 18–54), with MRI revealing structural abnormalities in 41% and EEG abnormalities in 44% (including focal and generalized epileptiform features). We also included 68 healthy controls (age 40.31 ± 12.84 [20–67]; sex 27/41 M/F), who were socioeconomically matched to the three epilepsy cohorts. As ALEC is not a conventional group analysis approach, it was important for us to capture a wide age range in the control cohort to create a reliable averaged template of the control population.

Full clinical, EEG, and MRI details for all participants are provided in Supplementary Table 1.

The study was approved by the Austin Health Human Research Ethics Committee (HREC/60011/Austin-2019 and HREC/68372/Austin-2022).

### Imaging parameters

MRI scanning was performed on a 3T Siemens PrismaFit at the Florey Institute of Neuroscience and Mental Health, Austin Campus, Melbourne, Australia. A 13:54-minute multi-band, multi-echo ^26,27^ echo-planar imaging sequence was acquired during movie watching (similar to ^28^).

Each of the three movies was interspersed with 20 seconds of rest, featuring a black screen with the word ‘Rest’ displayed in white font. The first movie was Inception, the second was The Social Network, and the last movie was Ocean’s Eleven.

fMRI parameters were a voxel size = 3×3×3 mm; TR = 0.9 s; TE = [15, 33.25, 51.5] ms; multiband factor = 4; GRAPPA = 2; field-of-view 216×216×132 mm; flip angle 30°; and an anterior–posterior phase-encoding. T1-weighted structural images were also acquired for the co-registration of fMRI data.

### Multi-echo fMRI preprocessing

We used fMRIPrep 23.1.2 ^29^, which is based on Nipype 1.6.1 ^30,31^. Below, we have used the fMRIPrep boilerplate as a template for our specific preprocessing strategy. A B0-nonuniformity map (or fieldmap) was estimated based on two (or more) echo-planar imaging references with topup ^32^. The T1-weighted image was corrected for intensity non-uniformity using N4BiasFieldCorrection ^33^, distributed with ANTs 2.3.3 ^34^, and was used as the T1w Reference throughout the workflow. The T1w Reference was then skull-stripped using a Nipype implementation of the antsBrainExtraction.sh workflow (from ANTs), with OASIS30ANTs as the target template.

Brain tissue segmentation of cerebrospinal fluid, white matter and grey matter was performed on the brain-extracted T1w using FSL fast ^35^. An anatomical T2w-reference map was computed after registration of 2 T2w images (after INU-correction) using mri_robust_template. Brain surfaces were extracted using FreeSurfer 7.4.0 before the fmriprep step, with recon-all ^36^. The previously estimated brain mask was refined using a custom variation of the method to reconcile ANTs-derived and FreeSurfer-derived segmentations of the cortical grey matter in Mindboggle ^37^. Volume-based spatial normalisation to standard space (FSL’s MNI ICBM 152 nonlinear 6^th^ Generation Asymmetric Average Brain Stereotaxic Registration Model ^38^) was performed through nonlinear registration with antsRegistration (ANTs 2.3.3), using brain-extracted versions of both T1w reference and the T1w template.

First, a reference volume and its skull-stripped version were generated by aligning and averaging 3 single-band references (SBRefs). Head-motion parameters with respect to the BOLD reference (transformation matrices and six corresponding rotation and translation parameters) are estimated before any spatiotemporal filtering using mcflirt ^39^. The estimated fieldmap was then aligned with rigid registration to the target echo-planar imaging reference run. The field coefficients were mapped onto the reference echo-planar imaging using the transform. BOLD runs were slice-time corrected to 0.4s (0.5 of the slice acquisition range 0s-0.8s) using 3dTshift from AFNI ^40^. A T2* map was estimated from the preprocessed echo-planar imaging echoes, by voxel-wise fitting the maximal number of echoes with reliable signal in that voxel to a monoexponential signal decay model with nonlinear regression. The T2*/S0 estimates from a log-linear regression fit were used for initial values.

The calculated T2* map was then used to optimally combine the preprocessed BOLD across echoes ^41^. The optimally combined time series was carried forward as the preprocessed BOLD.

The BOLD reference was then co-registered to the T1w reference using bbregister (FreeSurfer), which implements boundary-based registration ^42^. Co-registration was configured with six degrees of freedom. First, a reference volume and its skull-stripped version were generated using a custom methodology of fMRIPrep. Several confounding time-series were calculated based on the preprocessed BOLD: three region-wise global signals within the cerebrospinal fluid, the white matter, and the whole-brain masks. Gridded (volumetric) resamplings were performed using antsApplyTransforms (ANTs), configured with Lanczos interpolation to minimise the smoothing effects of other kernels ^43^. Non-gridded (surface) resamplings were performed using mri_vol2surf in FreeSurfer. Many internal operations of fMRIPrep use Nilearn 0.8.1 ^44^, mostly within the functional processing workflow.

After FreeSurfer and fMRIprep processing, six motion parameters (three translations and three rotations) were regressed out of the fMRI data, as well as three brain tissue signals (white matter, cerebrospinal fluid, and the whole brain signals), using fsl_glm. Then, bandpass filtering was performed with fslmaths on the residual fMRI data between frequencies of 0.01 and 0.1 Hz,.

### From Regional Homogeneity (ReHo) to Adjusted Local Estimates of Connectivity (ALEC)

Next, we calculated local connectivity via Regional Homogeneity (ReHo) ^17^ in the preprocessed and MNI space normalised fMRI data. ReHo is a voxel-based metric of brain local connectivity for fMRI data, which estimates the average correlation between a given voxel and its neighbouring voxels. ReHo is calculated using the Kendall tau rank correlation coefficient between a voxel and its nearest voxels within a surrounding 3D cube. The ReHo measure associated with *K* voxels inside a 3D cluster (*M* = 27) is computed as:

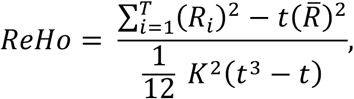

where *t* is the number of time points in the time series, *R_i_* is the sum rank of the *i^th^* time point over all neighbouring voxels within the cluster (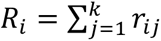 where *r_ij_* is the rank of the *i^th^* time point at the *j^th^* voxel) and *R̄* is the average of *R_i_*’s over time points. The metric ReHo always takes values from 0 (minimal local connectivity) to 1 (maximal local connectivity). ReHo maps were then smoothed with a Gaussian full-width-at-half-maximum kernel of 8 × 8 × 8 mm. Subject-specific grey matter masks were used for ALEC.

ALEC is a modified *z*-test, and its main statistical assumption is that the underlying data distribution is Gaussian with a mean of 0. We used a Rank-Based Inverse Normal Transformation ^45^ where ReHo feature voxels were first ranked and then transformed into a Gaussian shape with a distribution mean of 0 and a standard deviation of 1.

ALEC for voxel *i* is defined as:

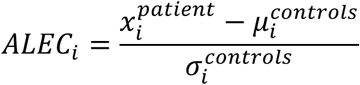

where *x_i_* the normalised ReHo voxel *i* of a single participant. *μ_i_* is the normalised mean and *σ_i_* is the standard deviation of ReHo voxel *i*, across 68 healthy controls.

### Statistical analysis

To minimise false-positive errors in voxel-wise ALEC maps, we applied a False Discovery Rate (FDR) ^46^ detection rate to statistically threshold voxel-wise ALEC maps at *p* < 0.05. In addition to individual voxel-wise ALEC maps, our primary outcomes in this study were i) descriptive voxel-wise ALEC maps summarising all subjects within each group; ii) whole-brain averaged ALEC per participant (where absolute mean values were used to capture both increased and decreased connectivity changes in the same metric); and iii) the log-transformed number of suprathreshold FDR-corrected ALEC voxels per participant, indexing the spatial extent of individual brain abnormalities. Outcomes ii) and iii), were examined with one-way ANOVA followed by Tukey Honestly Significant Difference (HSD) post hoc tests, *p* < 0.05, between DRE, new Dx and 1^st^ Sz groups. Associations between ALEC and age were assessed as epilepsy subjects had a wide range of ages (adults only), and to understand whether our results are related to ‘normal brain development’ in patients or reflect potential clinical changes. To do so, we were using Spearman’s *ρ* correlations within each epilepsy subgroup. Significance was set at *p* < 0.05 (two-tailed), with a multiple comparison correction using the Bonferroni method.

## Results

### Whole-brain average ALEC results

A one-way ANOVA revealed that whole-brain averaged ALEC was significantly different between groups, specifically DRE, new Dx, and 1^st^ Sz, *F*(2, 99) = 3.41, *p* = 0.037. Post hoc comparisons using the Tukey HSD (*p* < 0.05) test indicated that the mean score for the DRE group (M = 0.906, SD = 0.195) was greater than that of the new Dx (M = 0.810, SD = 0.134) and 1^st^ Sz (M = 0.811, SD = 0.186) groups– see Figure 1, left side.

**Figure 1:**
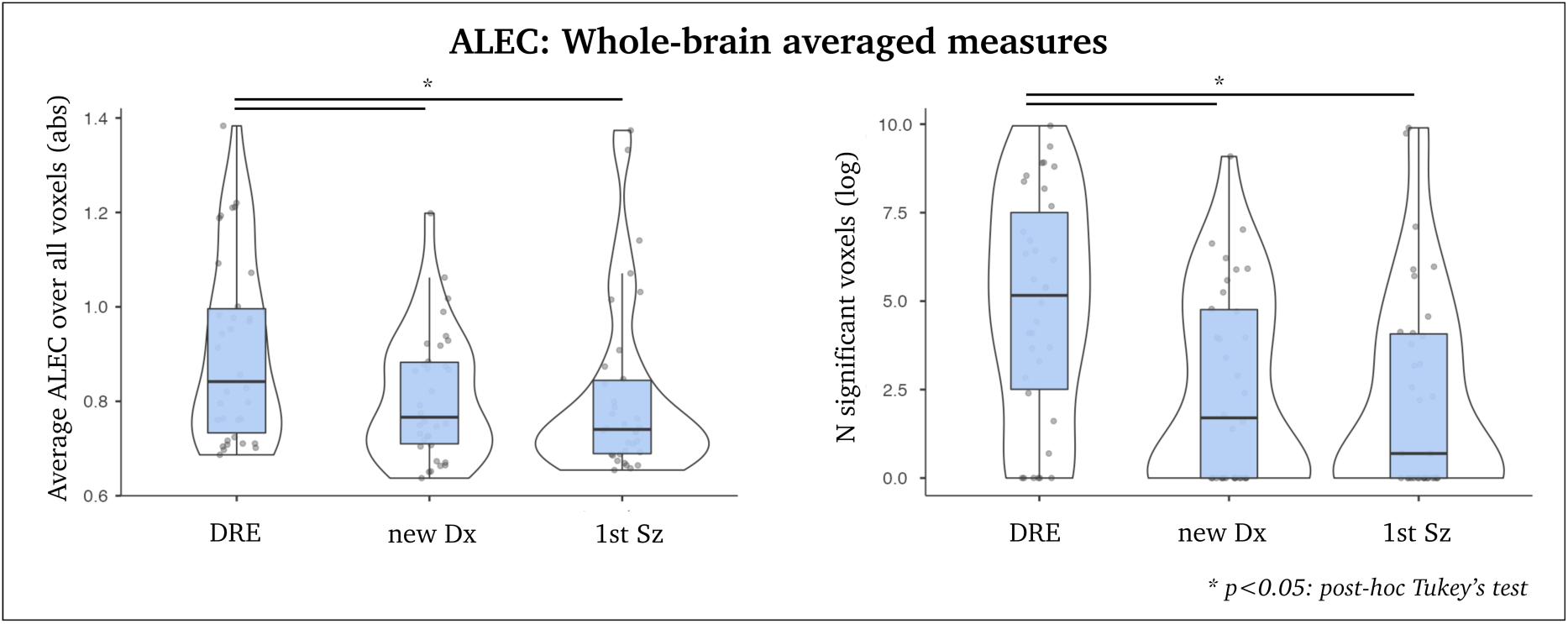
Summary of ALEC results, for average absolute ALEC values (increased and decreased connectivity) of the left side, and log-transformed number of significant voxels on the right side. Each dot denotes a single subject. The black line represents the median value, and the blue boxes represent the interquartile range.

A second one-way ANOVA also showed that the number of statistically significant ALEC voxels differed between groups, i.e., DRE, new Dx and 1^st^ Sz, *F*(2, 99) = 7.22, *p* < 0.001. Post hoc comparisons using the Tukey HSD (p < 0.05) test indicated that the mean score for the DRE group (M = 4.807, SD = 3.216) was greater than that of the new Dx (M = 2.542, SD = 2.744) and 1^st^ Sz (M = 2.345, SD = 2.925) groups. – see Figure 1, right side.

### Voxel-wise ALEC summation maps

Brain maps summarising all un-thresholded ALEC maps across individuals revealed elevated ALEC in the hippocampus, thalamus and brainstem, accompanied by widespread cortical decreases, in DRE (Figure 2A). In contrast, early-stage cohorts (new Dx and 1^st^ SZ) showed absent subcortical increases and fewer cortical decrements. See Figure 2B and C.

**Figure 2:**
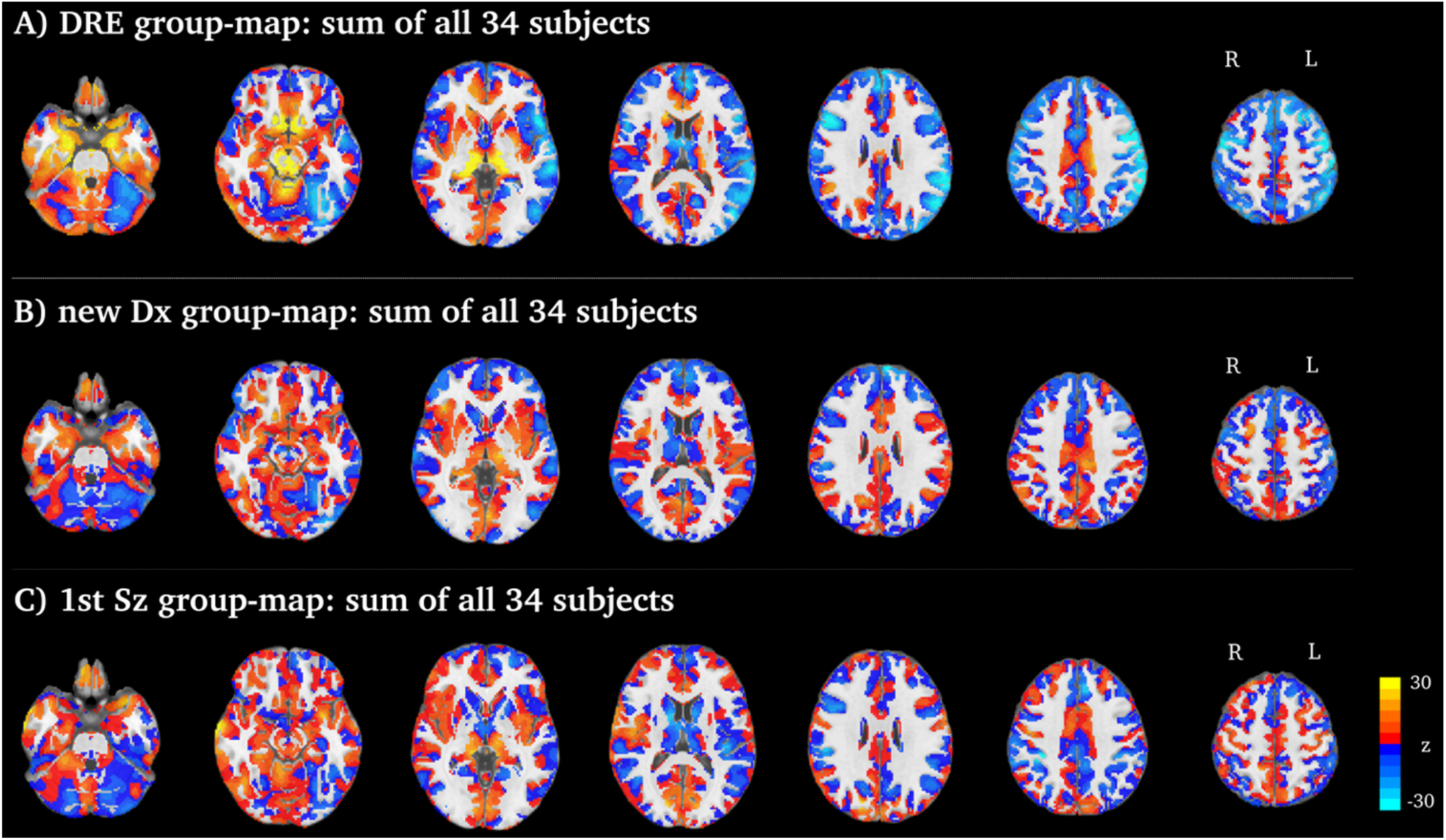
Summation of all un-thresholded voxel-wise ALEC maps. Hot colours indicate stronger connectivity, and cold colours indicate weaker connectivity, compared to controls.

A voxel-level ANOVA revealed no significant differences among the three groups after applying FDR correction. Given the absence of a voxel-wise group effect, we examined individual variability. We found that 11 out of 34 (32.4%) DRE subjects exhibited thalamic ALEC values at least one standard deviation above the control mean, whereas 8 out of 34 (23.5%) DRE subjects showed hippocampal ALEC values exceeding one standard deviation above the control mean.

These findings suggest that subcortical increases in DRE are likely driven by individual differences rather than being a consistent group-level effect.

### Correlation between ALEC and age

As seen in Figure 3, age correlated positively with whole-brain averaged ALEC (Spearman’s *ρ* = 0.723, *p* < 0.001) and number of significant voxels (Spearman’s *ρ* = 0.775, *p* < 0.001) in DRE, after Bonferroni correction. No statistically significant age–ALEC relationships were observed in the new Dx cohort (average ALEC - Spearman’s *ρ* = 0.051, *p* = 0.774; number of significant voxels - Spearman’s *ρ* = 0.054, *p* = 0.755) or 1^st^ Sz cohort (average ALEC - Spearman’s *ρ* = 0.163, *p* = 0.353; number of significant voxels - Spearman’s *ρ* = 0173, *p* = 0.327).

**Figure 3:**
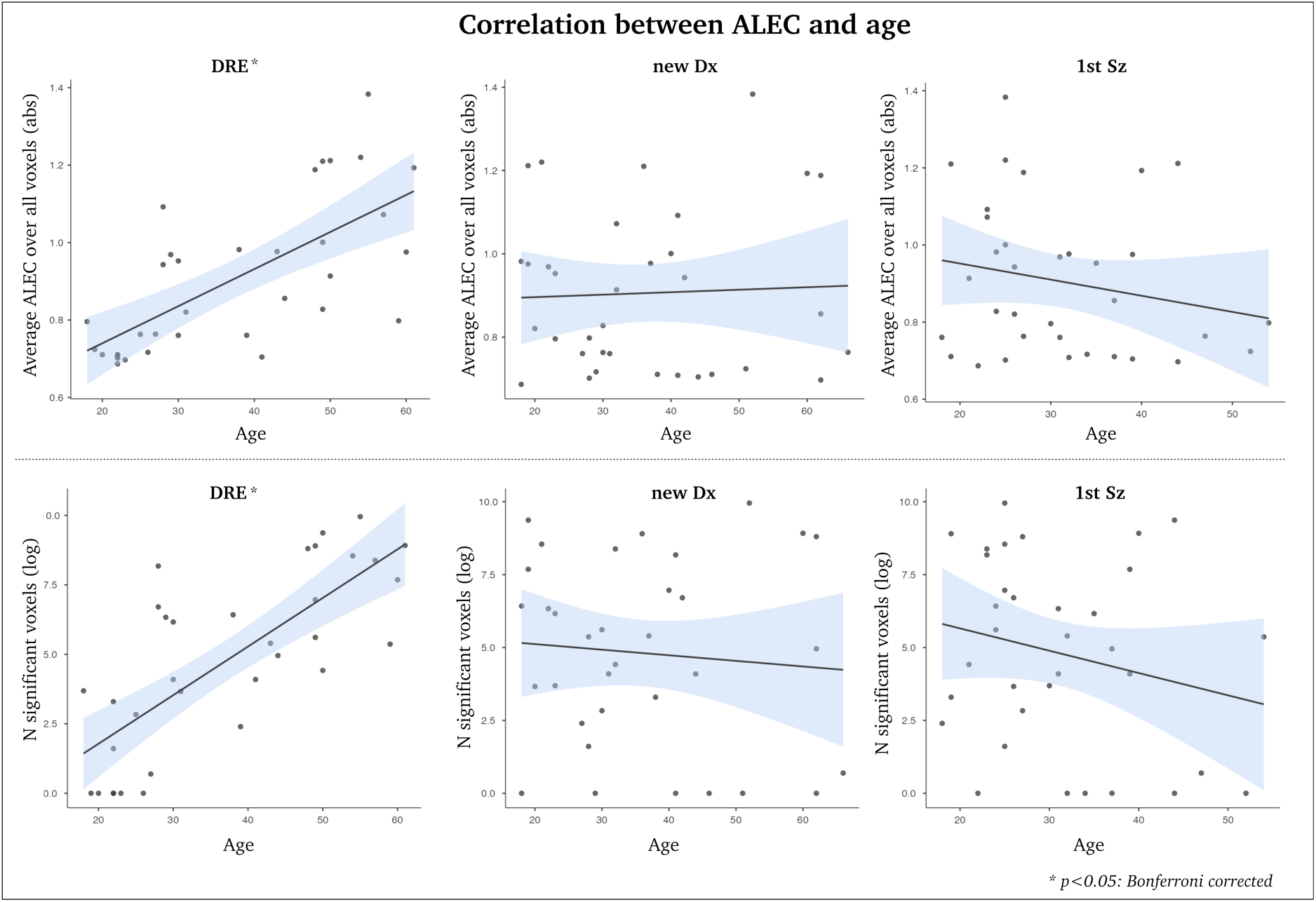
Scatterplots between ALEC metrics and age for DRE, new Dx and 1^st^ Sz (top = average ALEC; bottom = number of significant voxels). The black lines represent the best linear fit of the data, with the standard error as the shaded blue pattern.

### Correlation between ALEC and seizure duration in DRE

Next, we wanted to test whether ALEC was also correlated with seizure duration (years since epilepsy diagnosis), in DRE. We observed that seizure duration correlated positively with mean ALEC (Spearman’s *ρ* = 0.497, *p* = 0.002) and number of significant voxels (Spearman’s *ρ* = 0.472, *p* = 0.003) in DRE. These correlations were significant after Bonferroni correction. This suggests that the previous age-related correlation with ALEC is linked to seizure duration.

### ALEC case series in epilepsy

Next, we present six cases that showcase the clinical utility of individual ALEC maps. We present cases of drug resistance where ALEC helped identify potential pathologies and associated networks. These cases are visualised in Figure 5.

*Case #1:* The first case is a man in his sixties. Structural MRI shows evidence of left hippocampal sclerosis. EEG shows intermittent left temporal delta slowing, consistent with an underlying structural or functional abnormality. ALEC also showed increased connectivity in the hippocampus (left > right) and thalamus, together with decreased ALEC in the bilateral fronto-insular and cortical temporal lobes, as well as reduced connectivity in the anterior cingulate cortices and precuneus, all FDR corrected.

*Case #2:* This patient (a female in her fifties), has a previous left peri-amygdala lesion resection, sparing the hippocampus. Small-volume surgical margin gliosis, accompanied by minimal focal T2 hyperintensity within the CA3 aspect of the anterior left hippocampal head, raises the possibility of minimal focal hippocampal damage. EEG shows sharp discharges with some epileptiform features, commensurate with an active seizure disorder arising likely from a left hemisphere focus. Concordantly, ALEC showed increased connectivity in the ipsilateral hippocampus, with no other findings after FDR correction.

*Case #3:* This patient, in his forties, has a normal MRI and frequent interictal EEG epileptiform discharges, including focal slowing, over both right and left temporal regions independently. These findings are consistent with bilateral temporal lobe epilepsy. In alignment with the clinical findings, ALEC exhibits increased connectivity in the bilateral hippocampus and brainstem, as well as decreased connectivity in the cingulate cortex and left parietal lobe.

*Case #4:* This male patient (in the forties) shows with evidence of brain atrophy (parenchyma loss) in the parietal lobes. Otherwise normal MRI. Abnormal EEG showing left temporal focal delta and epileptiform sharp slow waves. ALEC shows ipsilateral decreased connectivity in the cortical temporal lobes and increased ipsilateral connectivity of the caudate, after FDR correction.

*Case #5:* A female in the twenties has a left bottom-of-the-sulcus dysplasia in the left precentral sulcus. Clinical features of focal motor features, but no definite abnormal EEG. There were no voxels surviving FDR correction in this patient, which is in line with our findings that ALEC tends to become more abnormal with age in DRE. However, there is a lateralised, sub-threshold increase of ALEC in the peri-lesional areas in the left precentral sulcus. This highlights the argument between clinical and statistical significance in this approach, a topic we revisit in the Discussion section.

*Case #6:* This man in his fifties has a previous right temporal lobectomy. Stable atrophy of the right hippocampus anteriorly. White matter hyperintensity suggestive of underlying chronic small vessel disease. EEG shows a correlate with right temporal epileptiform onset. ALEC exhibits extensive abnormalities, particularly increased connectivity in the ipsilateral piriform cortex, as well as in networks including the ventral striatum, brainstem, and thalamus.

Widespread decreases in ALEC are observed across cortical motor areas.

Above, we reviewed 8/34 (23.5%) of our participants with DRE. Beyond this, we used notes from clinical meetings, to perform a qualitative evaluation of clinical usefulness in the DRE cohort. In summary ALEC was helpful in 21/34 cases (61.7%), to aid further clinical evaluation. Our future outcome data is needed to fully confirm the clinical usefulness of this data. However, one of the main questions that arises at this point is whether ALEC can show indicators of patients at risk of developing DRE? Although we have limited outcome data at this point, we are presenting two patients (Case #7 from the 1^st^ SZ cohort and Case #8 from the new Dx cohort) who continue to experience seizures after the fMRI scan. These cases are visualised in Figure 6.

*Case #7:* This male in the twenties was scanned after his first seizure (i.e, 1^st^ Sz). Since he has had 4-5 impaired awareness seizures and a new diagnosis of epilepsy, likely temporal lobe epilepsy. He has a stable white matter disease, which is greater than expected for his age. No definitive epileptogenic abnormality is identified. EEG is normal. The patient has an extensive history of mental health problems, including previous suicide attempts. He experiences frequent visual, auditory and olfactory hallucinations. ALEC shows a widespread pattern of abnormalities, particularly increased connectivity in the bilateral thalamus and basal ganglia. Decreased connectivity is observed in the cortical temporal, parietal, and secondary visual cortical areas.

*Case #8:* This male (in the twenties) was scanned after her diagnosis of epilepsy (i.e., new Dx). Seizure diary shows at least 10 aware seizures within 6 months. MRI is normal, and the EEG shows an ictal rhythm over the right centro-parietal region (Cz, Pz > C4) around 10 seconds after clinical onset in most events. The patient experiences light-headedness and a sudden onset of dizziness, a sense of spinning, feeling heavy in the head, and the sensation that he is falling. Concordant with parietal lobule abnormalities, ALEC is increased in the ipsilateral superior parietal lobule and bilateral thalamus, after FDR correction.

## Discussion

This study introduces ALEC, a framework designed to detect patient-specific local connectivity deviations across clinical stages of epilepsy. Using ALEC, we identified increased local connectivity (Figure 1), with biologically coherent patterns in chronic epilepsy patients. Several patients displayed increased hippocampal and thalamic ALEC, as seen in individuals with DRE (see Figure 2, which summarises all ALEC maps). These are known epicentres for epilepsy initiation and propagation, and are not a surprising finding given that a substantial portion of DRE patients have various forms of temporal lobe epilepsy (see Supplementary Table 1).

Previous large-scale research from the ENIGMA project aligns with our findings, as it also demonstrates that atrophy of the hippocampus and thalamus, along with cortical areas, is associated with fMRI-based connectivity in temporal lobe epilepsy ^47^. Prior evidence also suggests that thalamic hyperexcitability may predict outcome after surgery ^48^, as well as its role in mediating seizures through deep brain stimulation ^49^. The accompanying cortical hypoconnectivity in our study may represent reduced local coherence in distributed association networks, a finding that is in line with previous research showing fMRI-based hypoconnectivity in temporal lobe epilepsy ^50,51^—cortical organisational shifts may be associated with a failure of network-based inhibitory control due to ongoing seizure activity ^52^.

Age and seizure duration are related to increases in ALEC (see Figures 3 and 4). This correlation suggests a progressive network disruption associated with epilepsy burden in DRE, as we did not observe a correlation between age and ALEC in new Dx or 1^st^ Sz cohorts. Together, this observation suggests that age is a clinical variable in our study, rather than reflecting normal age development across the lifespan. Epilepsy is known to accelerate brain ageing. Several studies have shown an increased rate of ageing across epilepsy subtypes ^53–56^. One of the challenges in this context is that our future aim is to attain early prediction of DRE, and brain abnormalities are likely more subtle during the earlier years of epilepsy. This raises the conundrum of the clinical versus statistical significance of ALEC, given that brain maps were often less significant in 1^st^ Sz and new Dx cohorts, as well as in younger people in the DRE cohort. Despite this challenge, we previously showed that functional brain connectivity changes can be detected within one year of a person’s first seizure, but only among individuals who continue to experience seizures after initiating anti-seizure medication. This suggests that recurrent seizures, rather than the effects of medication, may be a determinant of longitudinal changes in brain connectivity ^57^.

**Figure 4:**
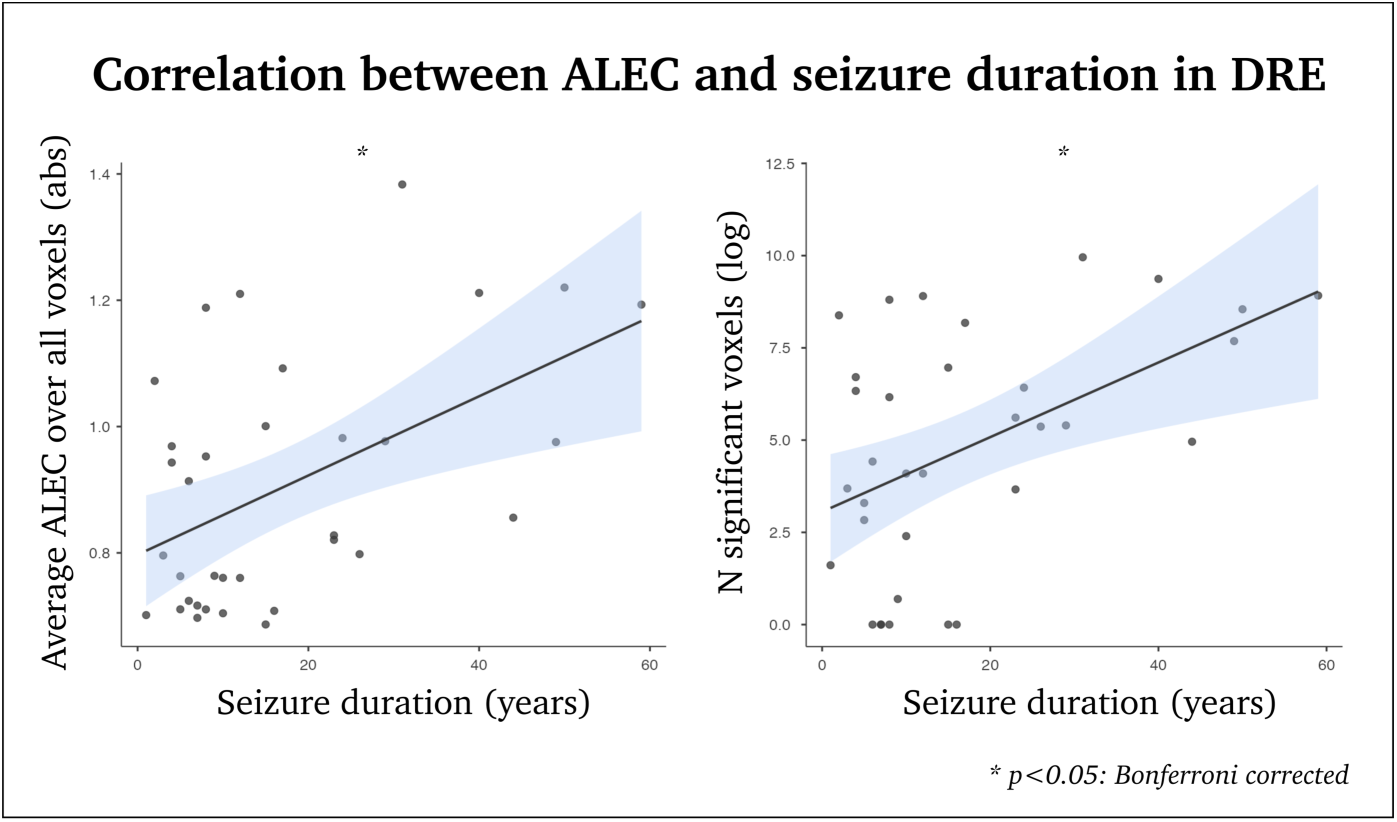
Scatterplots between ALEC metrics and seizure duration (years) for DRE. The black lines represent the best linear fit of the data, with a the standard error as a shaded blue pattern.

Group-level analyses often reveal subtle yet consistent abnormalities across patients. In contrast, individual-level metrics—such as ALEC—reflect large effect sizes with a higher potential of clinical relevance. This duality represents both the strength and the limitation of approaches like ALEC. Individual-based metrics yield rich, detailed information, underscoring the need for systematic frameworks to interpret these findings. Our clinical framework has evolved over the past decade through weekly multidisciplinary meetings, where advanced MRI results (e.g., ALEC, simultaneous EEG-fMRI, and language/motor fMRI mapping) are reviewed collaboratively by clinical and imaging experts. These discussions have led to consensus interpretations that provide clinically valuable insights into epilepsy-related network patterns and, in some cases, underlying pathology. For examples of cases where ALEC overlapped with known pathology, see Case #1 with hippocampal sclerosis and Case #5 with bottom-of-sulcus dysplasia, as shown in Figure 5. These findings also align with our previous observations, where excessive ReHo-based local connectivity identified a subtle epileptogenic lesion in an MRI-negative patient ^58^.

**Figure 5:**
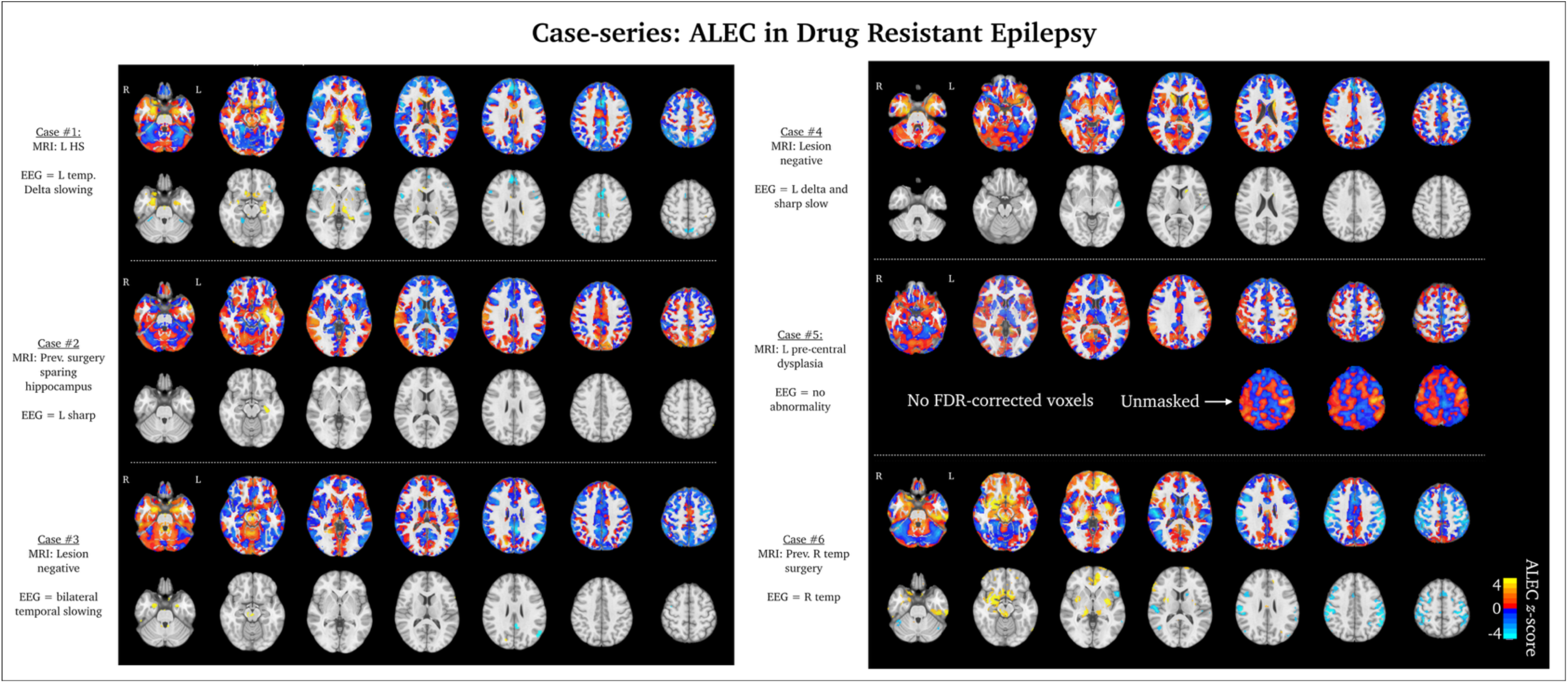
Individual brain maps of drug-resistant epilepsy. For each case, the top row represents unthresholded z-score ALEC maps, whereas the bottom row represents FDR-corrected ALEC maps. HS = hippocampal sclerosis, L = left, R = right. The colour scaling is the same for all brain maps (ALEC −4 to 4, which is a z-score compared to the control population).

One of the goals of the Australian Epilepsy Project is to leverage prospective data to predict individuals at risk of developing epilepsy. However, given the current absence of two-year outcome data for first-seizure and newly diagnosed epilepsy patients, we consider univariate statistical approaches—such as ALEC—as hypothesis-generating frameworks that help us explore and understand the underlying mechanisms of epilepsy. It remains to be seen whether the current ALEC results can be used in the future to predict epilepsy outcomes. However, our initial findings in people with recurrent seizures are showing significant ALEC alterations (Figure 6).

**Figure 6:**
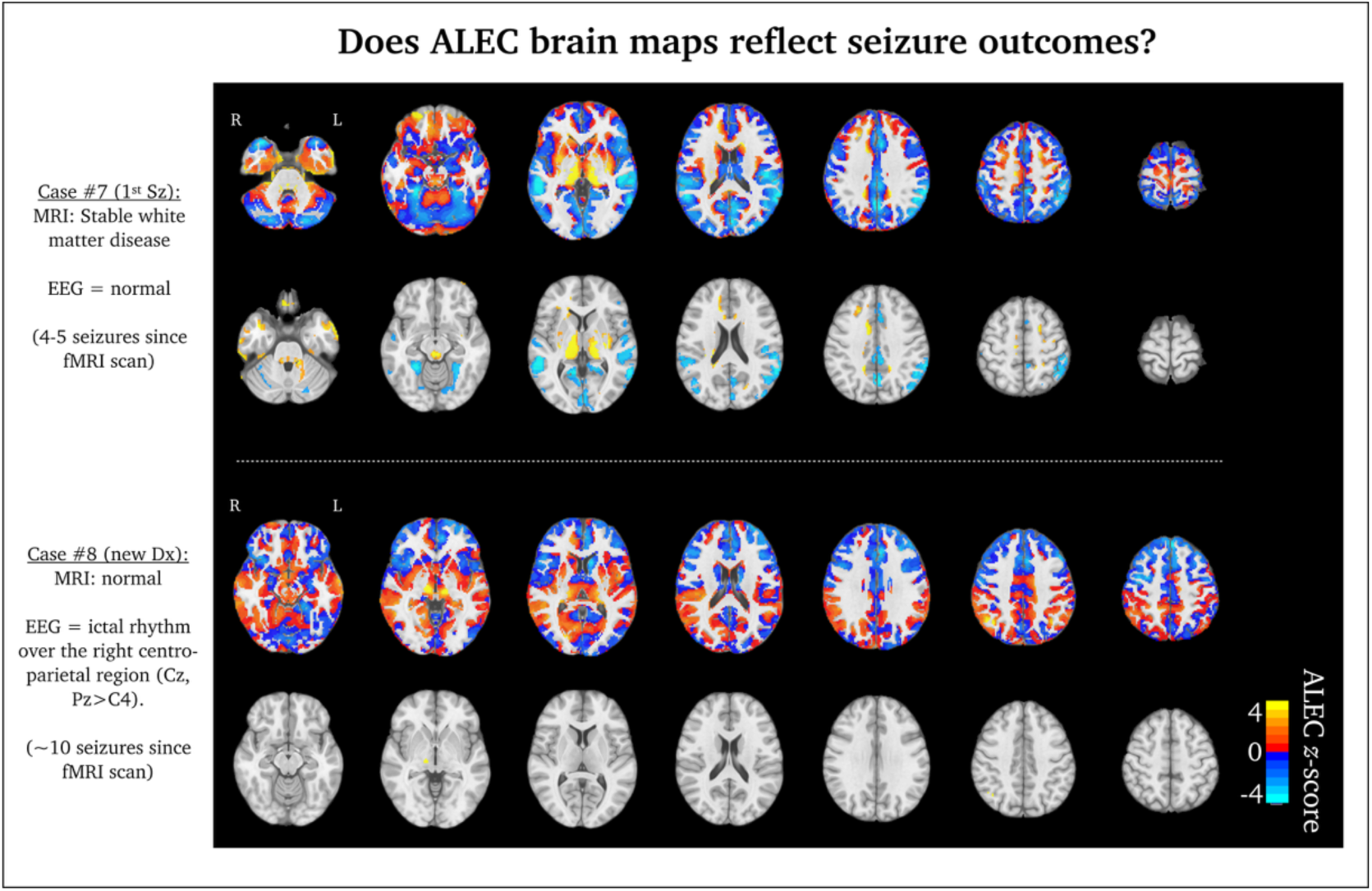
Individual brain maps for people with recurrent seizures, after the first scan. In each case, the top row represents un-thresholded z-score ALEC maps, whereas the bottom row represents FDR corrected ALEC maps. L = left, R = right. The colour scaling is the same for all brain maps (ALEC −4 to 4, which is a z-score compared to the control population).

Encouragingly, recent approaches have demonstrated that incorporating positron emission tomography (PET) imaging into structural imaging enhances the automated detection of epilepsy lesions, such as focal cortical dysplasia ^59^, using the machine learning Multi-centre Epilepsy Lesion Detection (MELD) approach ^60^. This suggests that incorporating functional neuroimaging into the existing framework may enhance the accuracy of automated epilepsy and lesion prediction. A related correlational observation in the literature has been the close relationship between ReHo and fluorodeoxyglucose-PET (FDG-PET), notably in default-mode and sensorimotor networks ^61–63^, potentially providing a new perspective on brain function that may be linked to the metabolic brain. In epilepsy, it is too early to postulate whether advanced fMRI can be used in conjunction with, or independently of, PET imaging. Future epilepsy studies with available fMRI and PET data in epilepsy are warranted. However, fMRI-based tools remain an attractive non-invasive investigation in the clinician’s toolbox.

To conclude, ALEC provides an individualised fMRI framework for detecting patient-specific connectivity abnormalities in epilepsy. Our findings reveal a pattern of local hyperconnectivity in drug-resistant cases, absent at early disease stages, that correlates with age and seizure duration. These results highlight ALEC’s potential as a precision imaging marker for stratifying drug resistance risk in epilepsy.

## Data availability

Note that we reported 8 of the 102 possible epilepsy cases in this study, across three cohorts. Given the abundance of potential information in this dataset, we are interested in collaborating and sharing ALEC data for further analysis and exploration. Please contact the Australian Epilepsy Project if you have any interest (https://www.epilepsyproject.org.au/contact-us). A MATLAB-based ALEC toolbox is available for download on GitHub (github.com/MangorPedersen/ALEC).

## Acknowledgements

We wish to thank Dr Jonas Haderlain and Dr Aaron Capon for their implementation of the ALEC pipeline in the Australian Epilepsy Project. We also want to thank all the clinicians and scientists in the Austin Hospital and Florey Institute CNRG program, who have provided valuable clinical insights into ALEC approaches over the past decade.

The Australian Epilepsy Project received funding from the Australian Government under the Medical Research Future Fund (Frontier Health and Medical Research Program, grant numbers MRFF75908 and RFRHPSI000008) and the Victoria State Government (Victorian-led Frontier Health and Medical Research Program). The Florey Institute of Neuroscience and Mental Health also acknowledges the strong support from the Victorian Government and, in particular, funding from the Operational Infrastructure Support Grant.

## The Australian Epilepsy Project investigators

David F. Abbott: Florey Institute of Neuroscience and Mental Health; conceptualization and funding acquisition. Subhaga Amarasekara: Florey Institute of Neuroscience and Mental Health; resources. Amanda Anderson: Florey Institute of Neuroscience and Mental Health; investigation, resources, funding acquisition. Rachel Hughes: Florey Institute of Neuroscience and Mental Health; project administration. Donna Hutchison: Florey Institute of Neuroscience and Mental Health; project administration. Paul Lightfoot: Florey Institute of Neuroscience and Mental Health; investigation, project administration. Saul Mullen: University of Melbourne; conceptualization and funding acquisition. Karen L. Oliver: University of Melbourne; conceptualization and funding acquisition. Heath R. Pardoe: Florey Institute of Neuroscience and Mental Health; conceptualization and funding acquisition. Mangor Pedersen: Auckland University of Technology; conceptualization and funding acquisition. Chris Tailby: Florey Institute of Neuroscience and Mental Health; conceptualization and funding acquisition. David N. Vaughan: Florey Institute of Neuroscience and Mental Health; conceptualization and funding acquisition. Anton De Weger: Florey Institute of Neuroscience and Mental Health; software, resources, data curation.

## Notes

### Competing Interest Statement

The authors have declared no competing interest.

