## Supplementary Table 1 for "Individualised Functional Brain Mapping Distinguishes Drug-Resistant from Early-Stage Epilepsy"

*Supplementary Table 1: Clinical characteristics of all participants in this study*

| Subject ID | Cohort | Gender | Age | MRI findings | EEG findings | Seizure duration |
|---|---|---|---|---|---|---|
| 001 | DRE | M | 40s | Possible cortical malformation in the superior left temporal gyrus | Normal | 10 years |
| 002 | DRE | F | 20s | No structural abnormality | Seizure and interictal discharges in the parietal lobe | 8 years. |
| 003 | DRE | M | 60s | No structural abnormality | Slow activity in the left mid temporal region. | 49 years. |
| 004 | DRE | M | 20s | Incidental finding: arachnoid cyst | Left sided sz focus. | 17 years. |
| 005 | DRE | F | 50s | DNET/ganglioglioma right intra-axial mesial temporal lobe | Normal | 2 years. |
| **006 (case 6)** | DRE | F | 50s | Minor resection margin gliosis at the posterior aspect of the right anterior temporal lobectomy. Atrophy of the right hippocampus anteriorly which may also be postoperative. | Right temporal epileptiform activity. | 40 years. |
| **007 (case 5)** | DRE | M | 20s | Bottom of the sulcus dysplasia involving the left precentral sulcus | 13 motor seizures, clinical features lateralise to the posterior aspect of the left frontal lobe. | 16 years. |
| 008 | DRE | M | 20s | No structural abnormality | Normal | 4 years. |
| 009 | DRE | M | 40s | Prior right fronto-temporal craniotomy with a small region of cortical and subcortical atrophy involving | Abnormal awake EEG with occasional right temporal theta slow waves and sharp transients. | 8 years. |
| 010 | DRE | F | 20s | No structural abnormality | Generalised sharp-and-slow-waves and polyspike and slow waves were observed with right frontal dominance. | 7 years. |
| 011 | DRE | M | 30s | 7mm left anterior temporal pole encephalocele with suspected associated gliosis | Frequent left temporal sharp waves. One event recorded with focal onset in the left hemisphere. | 8 years. |
| **012 (case 2)** | DRE | M | 50s | Prior left anterolateral temporal lobe resection sparing the left hippocampus. Minimal focal T2 hyperintensity within the anterior left hippocampal head. | Left structural or functional abnormality. | 26 years. |
| 013 | DRE | F | 20s | Right hippocampal sclerosis. Signal abnormality in the left amygdala suspicious for a developmental neoplasm. | Right temporal lobe epilepsy with rapid propagation of the contralateral hemisphere. | 4 years. |
| **014 (case 3)** | DRE | F | 40s | No structural abnormality | Frequent interictal epileptiform discharges over both right and left temporal regions independently. | 15 years. |
| 015 | DRE | F | 50s | Possible prior cytotoxic lesion of the corpus callosum | Numerous bilateral independent interictal discharges from right and left anterior quadrants. R>L. | 6 years. |
| **016 (case 4)** | DRE | F | 40s | No structural abnormality | Abnormal EEG showing left temporal focal delta and epileptiform sharp slow. | 29 years. |
| 017 | DRE | M | 30s | Unusual gyral configuration in the medial left parieto-occipital region is developmental and could be dysplastic. | Not available | 10 years. |

| | | | | | | |
|---|---|---|---|---|---|---|
| **018 (case 1)** | DRE | F | 60s | Left hippocampal sclerosis | Seizure captured arising from the left temporal region. | 59 years. |
| 019 | DRE | F | 50s | Incidental finding: Microhaemorrhage in the right occipital lobe | Seizure arising from the left anterior temporal area. | 31 years. |
| 020 | DRE | F | 30s | Probable small right temporal pole encephalocele | Rare left temporal sharps. | 24 years. |
| 021 | DRE | M | 10s | No structural abnormality | Normal | 6 years. |
| 022 | DRE | F | 10s | No structural abnormality | Normal | 3 years. |
| 023 | DRE | M | 20s | No structural abnormality | Underlying structural or functional pathology at the right temporal/parietal region. | 1 year. |
| 024 | DRE | F | 30s | Left frontal and occipital lesions: neurocysticercosis is a likely cause | All seizures arose from the left anterior hemisphere (F3 predominantly, with Cz and T3 involved). | 23 years. |
| 025 | DRE | F | 50s | Evidence of previous bifrontal and bitemporal electrode implantation. Features compatible with anteromedial right superior frontal gyrus focal cortical dysplasia (Type 2b). | Nocturnal frontal lobe epilepsy with a right frontal focus. | 50 years. |
| 026 | DRE | F | 20s | Incidental finding: left small developmental venous anomaly | EEG changes arising from the left temporal region. Consistent with a clinical diagnosis of left temporal lobe epilepsy. | 5 years. |
| 027 | DRE | F | 20s | Incidental finding: pineal cyst | frontotemporal slowing is suggestive of left frontotemporal structural abnormality or dysfunction | 9 years. |
| 028 | DRE | M | 40s | Previous left inferior frontal gyrus cavity. | focal interictal discharges of spike and wave were seen over left frontal region. no event was captured. The findings were consistent with diagnosis of left frontal lobe epilepsy | 44 years. |
| 029 | DRE | M | 30s | Superior left Amygdala T2 hyperintense nodules – possibility of a DNET/ganglioglioma | Occasional left temporal sharp and slow waves. | 12 years. |
| 030 | DRE | M | 40s | No structural abnormality | R temporal sharp waves during sleep | 23 years |
| 031 | DRE | M | 20s | No structural abnormality | Clinical and EEG features indicate a left frontal focus | 15 years. |
| 032 | DRE | F | 40s | Asymmetric hippocampal shape and heterogeneous T2 signal and HIMAL | Not available | 12 years. |
| 033 | DRE | M | 20s | No structural abnormality | right posterior quadrant and bitemporal slowing consistent with underlying functional/ structural abnormalities. | 5 years. |
| 034 | DRE | F | 20s | No structural abnormality | One seizure was captured which could be lateralized to the left. | 7 years. |
| 100 | 1$^{st}$ Sz | M | 40s | Small volume chronic micro haemorrhage and associated gliosis and small volume adjacent encephalomalacia within the right median cerebellar hemisphere. | Normal | |
| 101 | 1$^{st}$ Sz | F | 40s | No structural abnormality | Normal | |

| | | | | | |
|---|---|---|---|---|---|
| 102 | 1st Sz | M | 10s | No structural abnormality | Normal |
| 103 | 1st Sz | F | 40s | No structural abnormality | Normal |
| 104 | 1st Sz | M | 30s | No structural abnormality | Normal |
| 105 | 1st Sz | F | 10s | No structural abnormality | Normal |
| 106 | 1st Sz | M | 40s | Incidental finding: Generalised parenchymal volume reduction | Not available |
| 107 | 1st Sz | F | 40s | No structural abnormality | Normal |
| 108 | 1st Sz | F | 60s | Incidental: Nonspecific white matter signal hyperintensities are more extensive than expected for age. | Normal |
| 109 | 1st Sz | F | 60s | Several meningiomas. The larger lesions in the left frontal and right parietal region abut the adjacent cortex | Normal |
| 110 | 1st Sz | F | 20s | No structural abnormality | Normal |
| **111 (case 7)** | 1st Sz | M | 20s | Incidental finding: Stable white matter disease, greater than expected for age | Normal |
| 112 | 1st Sz | F | 20s | No structural abnormality | Normal |
| 113 | 1st Sz | M | 40s | No structural abnormality | Normal |
| 114 | 1st Sz | F | 30s | right frontal lesion is most compatible with a meningioma | Normal |
| 115 | 1st Sz | F | 30s | Inferolateral posterior left temporal encephalocele with possible gliosis, Multiple other bilateral probable encephaloceles | Normal |
| 116 | 1st Sz | M | 20s | left central sulcus focal cortical dysplasia | Not available |
| 117 | 1st Sz | M | 60s | Incidental finding: moderate burden of microvascular ischaemia | Normal |
| 118 | 1st Sz | M | 50s | No structural abnormality | Normal |
| 119 | 1st Sz | M | 10s | No structural abnormality | Normal |
| 120 | 1st Sz | F | 50s | No structural abnormality | Not available |
| 121 | 1st Sz | M | 20s | Possible dysplastic abnormality in the left precentral gyrus. | Abnormal EEG showing generalised spike wave discharge bursts spontaneously. The findings are consistent with a genetic generalised epilepsy syndrome. |
| 122 | 1st Sz | F | 20s | No structural abnormality | Normal |
| 123 | 1st Sz | M | 20s | No structural abnormality | Normal |
| 124 | 1st Sz | M | 20s | Tiny irregularities at the inferior aspect of the right frontal pole | Abnormal EEG recording suggestive of right hemispheric epileptogenicity with underlying structural abnormality or dysfunction and mild encephalopathy. |
| 125 | 1st Sz | F | 30s | A large developmental venous anomaly drains left frontal white matter and basal ganglia to the left insular region, hippocampal fissure cysts | Normal |

| | | | | | |
|---|---|---|---|---|---|
| 126 | 1st Sz | M | 60s | No structural abnormality | Normal |
| 127 | 1st Sz | M | 60s | Incidental finding: Parenchymal loss in the parietal lobes is greater than expected for age | Normal |
| 128 | 1st Sz | F | 30s | No structural abnormality | Abnormal EEG with a robust photoparoxysmal response. This EEG finding is associated with idiopathic generalised Epilepsy. |
| 129 | 1st Sz | M | 30s | No structural abnormality | Normal |
| 130 | 1st Sz | F | 10s | No structural abnormality | Normal |
| 131 | 1st Sz | M | 30s | No structural abnormality | Normal |
| 132 | 1st Sz | M | 30s | No structural abnormality | Normal |
| 133 | 1st Sz | M | 20s | No structural abnormality | Normal |
| 200 | New Dx | M | 30s | Tiny cluster of left anterior temporal pole 1-2mm encephaloceles | There are sharp waves in the R temporal region most prominent at T4/T6. On one occasion, there was suspicion for a left temporal sharp wave. |
| 201 | New Dx | F | 30s | No structural abnormality | Normal |
| 202 | New Dx | F | 30s | Left lateral temporal lobe Caveroma, Giant perivascular space within the right cerebral crus. | Normal |
| 203 | New Dx | F | 20s | No structural abnormality | Abnormal prolonged interictal recording, with generalised epileptiform discharges recorded, in keeping with an underlying idiopathic generalised epilepsy syndrome |
| 204 | New Dx | M | 20s | Incidental finding: White matter signal hyperintensities are more extensive than expected for age with a single left centrum semiovale lesion measuring up to 6 mm in diameter | Abnormal awake and drowsy EEG showing left anterior temporal sharp-slow wave discharges and intermittent focal delta slowing in keeping with left temporal lobe epilepsy |
| 205 | New Dx | M | 40s | Absence of the septum pellucidum with extensive bilateral peri-sylvian predominant polymicrogyria, most in keeping with underlying septo-optic dysplasia spectrum. | Normal |
| 206 | New Dx | M | 30s | No structural abnormality | Normal |
| 207 | New Dx | M | 20s | Amorphous white matter signal extending between the anterolateral left inferior and middle frontal gyri/frontal operculum and frontal horn of the left lateral ventricle most likely reflect presence of a left frontal cortical dysplasia (type 1 or 2a) or possibly MOGHE (mild malformation of cortical development with oligodendroglial hyperplasia and epilepsy). | Not available |
| 208 | New Dx | F | 20s | Left frontal trans-mantle vascular structures likely reflecting venous angiomas | Normal |

| | | | | | |
|---|---|---|---|---|---|
| 209 | New Dx | M | 40s | Small bilateral inferior temporal encephaloceles. | The background is slowed bi-frontally with frequent single spike wave and polyspike wave discharges captured throughout study. No electronical seizures captured. These findings are most consistent with a diagnosis of idiopathic (genetic) generalised epilepsy |
| 210 | New Dx | M | 30s | No structural abnormality | Normal |
| 211 | New Dx | M | 50s | No structural abnormality | Normal |
| 212 | New Dx | M | 30s | 5 cm left frontal cystic lesion, Small 5 mm right anterolateral temporal encephalocele, with impression of minor parenchymal gliosis | Normal |
| 213 | New Dx | F | 20s | Incidental finding: Right middle cranial fossa arachnoid cyst. | Normal |
| 214 | New Dx | F | 20s | Incompletely characterised 5 mm fourth ventricular nodule abutting the dorsal midbrain and inferior aspect of the fourth ventricular choroid plexus, immediately cranial to the level of foramen of Magendie. Tiny, 1-2mm, left anterior temporal encephaloceles | Normal |
| 215 | New Dx | F | 30s | No structural abnormality | Normal |
| 216 | New Dx | F | 10s | 5.5 mm unilateral right thalamic T2 FLAIR hyperintense focus is nonspecific | Normal |
| 217 | New Dx | F | 40s | large left frontal cavernoma | Abnormal EEG due to the observed left frontotemporal (F7T3) rhythmic patterns - the significance of this is not entirely clear, and an active seizure focus cannot be excluded |
| **218 (case 8)** | New Dx | M | 20s | No structural abnormality | EEG shows an ictal rhythm over the right centro-parietal region (Cz, Pz > C4) around 10 seconds after clinical onset in most events |
| 219 | New Dx | M | 20s | 15 mm region of cortically based abnormality involving the posterior inferior right temporal lobe | Abnormal routine awake and drowsing EEG to presence of grade III photoparoxysmal response |
| 220 | New Dx | M | 50s | Incidental finding: Generalised parenchymal volume reduction. | Normal |
| 221 | New Dx | F | 30s | No structural abnormality | Abnormal EEG due to a robust, repeatable photoparoxysmal response. This would be in keeping with a photosensitive idiopathic generalised epilepsy |
| 222 | New Dx | F | 20s | solid and cystic cortically based left superior temporal gyrus neoplasm | Abnormal awake and drowsy EEG due to persistent, fast beta activity over the left temporal lobe at T3 |

| | | | | | |
|---|---|---|---|---|---|
| 223 | New Dx | M | 20s | 4 mm right anterior temporal pole encephalocele without associated gliosis | Normal |
| 224 | New Dx | M | 20s | 5 mm right anterior inferior temporal encephalocele with possible gliosis | Normal |
| 225 | New Dx | M | 10s | No structural abnormality | A mildly abnormal EEG showing occasional left temporal delta slowing as well as occasional diffuse polymorphic theta-delta in drowsing. The findings are non-diagnostic |
| 226 | New Dx | M | 40s | No structural abnormality | Normal |
| 227 | New Dx | F | 30s | Incidental finding: Non-specific white matter signal hyperintensities are more extensive than expected for age | Bilateral independent anterior quadrant changes seem perhaps most frequently on the left |
| 228 | New Dx | F | 30s | tiny (1-2 mm) left anterior inferior temporal encephaloceles | Normal |
| 229 | New Dx | M | 20s | No structural abnormality | Abnormal EEG due to intermittent right temporal polymorphic slowing, consistent with a structural or functional abnormality. No definite epileptiform abnormalities. |
| 230 | New Dx | M | 20s | No structural abnormality | The record is unstable with a marked excess of disorganised theta in all areas which is probably inter-ictal in nature. |
| 231 | New Dx | F | 10s | Irregular gyral pattern involving both temporal lobes anterolaterally is most compatible with dysgyria | Abnormal awake EEG due to grade 3-4 photoparoxysmal response. This finding can occur in a patient with a diagnosis of GGE |
| 232 | New Dx | M | 20s | No structural abnormality | Normal |
| 233 | New Dx | M | 30s | No structural abnormality | Normal |
